# Antibiotic prescribing in remote versus face-to-face consultations for acute respiratory infections in English primary care: An observational study using TMLE

**DOI:** 10.1101/2023.03.20.23287466

**Authors:** Emma Vestesson, Kaat De Corte, Paul Chappell, Elizabeth Crellin, Geraldine M. Clarke

## Abstract

**Background:** The COVID-19 pandemic has led to an ongoing increase in the use of remote consultations in general practice in England. Though the evidence is limited, there are concerns that the increase in remote consultations could lead to more antibiotic prescribing.

**Methods:** We used patient-level primary care data from the Clinical Practice Research Datalink to estimate the association between consultation mode (remote vs face-to-face) and antibiotic prescribing in England for acute respiratory infections (ARI) between April 2021 – March 2022. We used targeted maximum likelihood estimation, a causal machine learning method with adjustment for patient-, clinician- and practice-level factors.

**Findings:** There were 45,997 ARI consultations (34,555 unique patients), of which 28,127 were remote and 17,870 face-to-face. For children, 48% of consultations were remote whereas for adults 66% were remote. For children, 42% of remote and 43% face-to-face consultations led to an antibiotic prescription; the equivalent in adults was 52% of remote and 42% face-to-face. Adults with a remote consultation had 23% (Odds Ratio (OR) 1.23 95% Confidence Interval (CI): 1.18-1.29) higher chance of being prescribed antibiotics compared to if they had been seen face-to-face. We found no significant association between consultation mode and antibiotic prescribing in children (OR 1·04 95% CI 0·98-1·11).

**Interpretation:** This study uses rich patient-level data and robust statistical methods and represents an important contribution to the evidence base on antibiotic prescribing in post-COVID primary care. The higher rates of antibiotic prescribing in remote consultations for adults are cause for concern. We see no significant difference in antibiotic prescribing between consultation mode for children. These findings should inform antimicrobial stewardship activities for health care professionals and policy makers. Future research should examine differences in guideline-compliance between remote and face-to-face consultations to understand the factors driving antibiotic prescribing in different consultation modes.

**Funding:** No external funding.

**Research in context:** *Evidence before this study:* Use of remote consultations in general practice has increased rapidly since the onset of the COVID-19 pandemic. Concerns have been raised that antibiotic prescribing rates may be higher in remote compared with face-to-face consultations. Acute respiratory infection (ARI) is the most common reason for an antibiotic prescription in adults making it one of the most important areas of prescription practice for antibiotic use. Empirical studies investigating the differences in antibiotic prescribing rates between online and remote consultations have produced mixed findings, in general and for ARIs specifically. Recent review-type articles on the topic - including a 2020 qualitative systematic review and a 2021 meta-analytic systematic review – have reported mixed results when comparing online and face-to-face consultations with some showing higher and others lower antibiotic prescribing in remote consultations. Furthermore, many of the studies that were included in the reviews were at risk of bias due to a failure to control for demographic and clinical differences between patients in remote versus face-to-face consultations.

*Added value of this study:* This is the first England wide study estimating the difference in antibiotic prescribing between consultations modes in the post-covid setting where remote consultations are as common as face-to-face consultations. It is also the first study in this setting to apply TMLE – doubly robust causal machine learning method. We found that an adult was 23% more likely to be prescribed an antibiotic for an ARI in a remote compared with a face-to-face consultation with a general practitioner in England. There was no evidence for a difference in children. Our findings are based on an analysis of a representative sample of almost 46,000 GP consultations for ARIs in general practice in England and controls for patient-, clinician- and practice-level factors that are associated with both consultation mode and with antibiotic prescribing. As such, our findings are at a smaller risk of bias from unobserved confounding than the previous research examining this issue and therefore represent an important contribution to the evidence base.

*Implications of the available evidence:* Taken together with the existing body of evidence on this topic, our results showing higher prescribing in remote consultations are cause for concern. The factors affecting antibiotic prescribing and the interaction with consultation mode are complex and will require further research to unpick. The existing evidence including this study have largely focused on prescribing rates, and do not investigate the appropriateness of antibiotics prescribing in remote compared to face-to-face consultations. Further investigation is required to explain the discrepancy between consultation modes. The growing body of evidence in this area has relevance for future antimicrobial stewardship activities and should be used to inform the ongoing development of antibiotic prescribing guidelines for remote consultations.

## Introduction

The development of antibiotic resistance is a global public health issue largely fuelled by antibiotic overprescribing. Most antibiotic prescribing in England happens in general practice: 72·1% of total antibiotic prescribing in 2021 (1) of which approximately 20% is inappropriate. (2) This makes general practice an important focus of antibiotic stewardship activities. The digitalisation of general practice was accelerated by the COVID-19 pandemic and has led to an ongoing increase in the use of remote consultations for all ages. (3) The full extent of the consequences of this rapid shift towards ‘telemedicine’ is still unclear and one area of concern is increased antibiotic prescribing in remote consultations. A survey of GPs in the UK reported that 67% of GPs thought that telehealth has increased their antibiotic prescribing to either a great extent or some extent which corroborates this concern.(4) As a GP cannot observe or examine the patient in a remote consultation in the same way as in a face-to-face consultation, it has been hypothesised that GPs might increase antibiotic prescribing to be ‘on the safe side’. (5)

Acute respiratory infections (ARIs) account for the greatest number of antibiotic prescriptions in UK general practice. (6) Remote consultations for ARIs could have a role in reducing spread of infection but over-prescription is a particular risk with these conditions. This is because they are often self-limiting and/or commonly caused by viruses rather than bacteria, (7) and because patients with cold or sore throat symptoms often request or even pressure GPs for antibiotics. (8)

There is limited evidence on the differences in ARI antibiotic prescribing between patients seen face-to-face and those seen remotely - in particular, in a post-pandemic setting where close to 50% of consultations are remote. Some observational studies have shown increases in prescribing in remote consultations for certain ARIs (9,10) but other analyses have found a decrease in prescribing in remote consultations (9,11) or no difference. (10,12) Additionally, a 2021 meta-analytic systematic review (13) and a 2020 qualitative systematic review (14) of the impact of remote consultation on antibiotic prescribing both proved inconclusive. Additionally, the majority of the relevant primary studies (9,11,12,15) and systematic reviews are at risk of bias due to a failure to control for demographic and clinical differences between patients who are seen remotely and face-to-face. Only one primary study in children used matching to adjust for baseline differences in covariates. (10) A more comprehensive understanding of differences in antibiotic prescribing in remote versus face-to-face ARI consultations is required, to inform remote consultation and antibiotic stewardship policy going forward.

In this study, we compare antibiotic prescribing in patients that were seen remotely by a GP for an ARI compared to patients that were seen face-to-face or using a mix of face-to-face and remote consultations. Analyses were conducted separately for adults and children under 16.

## Methods

### Study design and data

We performed a cohort study using person-level data from the Clinical Practice Research Datalink (CPRD) Aurum between January 2018 and July 2022. CPRD Aurum is a database with routinely collected data from primary care practices that uses EMIS Web®. CPRD contains data for over 40 million patients from 1,332 practices in England as of May 2022. Patients are broadly representative of the English population based on age, sex, and deprivation. CPRD also linked their data to the 2015 indices of multiple deprivation (IMD) at the patient-level, and to the 2011 urban-rural classification at the practice-level. The study protocol was approved by CPRD’s Research Data Governance (protocol number: 21_000357). The data was linked to the ONS infection survey by region (23 September 2022 release).

### Cohort eligibility criteria

We studied patients registered at general practices that participated in CPRD Aurum. Eligibility criteria were applied at both practice- and patient-level. 400 practices in England were sampled at random. Eligible patients were those with acceptable data quality (verified by CPRD); registered at one of the 400 practices at any point between January 2018 and March 2021; recorded as either male or female sex; and eligible for area-level linkage to the index of multiple deprivation (IMD). 600,000 patients were then randomly sampled from the eligible patients. Three GP practices were further identified by CPRD as having duplication issues and were excluded from the analysis.

### Analysis dataset

Consultations for acute respiratory infections were identified (see supplementary materials for codelists). This list was based on previously published lists of read codes. (16) There were five subgroups of ARIs: lower respiratory tract infections (LRTI), upper respiratory tract infections (URTI), sinusitis, otitis externa and otitis media as well as COVID. A consultation could be coded as more than one infection subtype.

Only consultations carried out by GPs were included. GPs are likely to be responsible for most of the antibiotic prescribing for ARIs in general practice and have a higher proportion of remote consultations than other health care professionals. There were 67,324 ARI consultations carried out by GPs between 1 April 2021 and 22 March 2022. This period spans the removal of the stay-at-home order to the most recent available data. ARI consultation records for the same patient happening on the same day were grouped together as these were likely to be part of the same consultation (49,451). ARI consultations happening in a 7-day period were grouped together retaining the date of first consultation. If the consultation mode was the same for all grouped consultations, it was coded as such; if there was a mix of face-to-face and remote consultations, it was recorded as face-to-face (henceforth referred to as mixed consultations). If antibiotics were prescribed in any of the grouped consultations, the prescription was retained in the analysis dataset.

### Variable Selection

To optimize the potential for variable adjustment as well as to ensure the exchangeability at baseline of the treatment arms, as many characteristics as possible were included in the dataset at several covariate levels: consultation-, patient-, clinician-, and practice-level. We adjusted for factors (or their proxies) known to be associated with antibiotic prescribing. These included comorbidities such as asthma or COPD, (17–20) health need, rurality, ethnicity, deprivation, (21) region (22), CCG, (23) clinician characteristics, (20,24) and overall practice consultation rates based on all patients in the sample.

We also adjusted for variables that were identified by experts to be associated with either antibiotic prescribing or having a remote consultation. These included COVID-19 infection in the last 7, 30 or 365 days before the consultation, number of remote or face-to-face consultations in the last 7, 30 or 365 days before the consultation and regional ONS COVID-19 infection prevalence.

All covariates were included in both the treatment and outcome models.

### Identification

To identify the treatment effect of consultation mode on antibiotic prescribing, we made the following causal assumptions. The stable unit treatment value assumption, i.e., the treatment status of any individual did not affect the potential outcomes of other individuals. This likely holds as one patient having a remote consultation is unlikely to have an impact on the decision to prescribe antibiotics to a different patient. The second assumption was no unmeasured confounding. In other words, the exposure mechanism and potential outcomes were independent after conditioning on our defined set of covariates. Thirdly, we assumed positivity, i.e., within strata of the set of covariates, all consultations had a nonzero probability of receiving either exposure condition. By design, only patients with a respiratory infection were included and those can be seen both remotely or face-to-face unlike consultations for other purposes such as vaccinations or wound management. We checked minimum and maximum of the propensity scores from the treatment model for violations of the positivity assumption.

We defined the statistical target parameters as the average treatment effect (ATE) and the odds ratio.

### Validation of exchangeability assumption

The standardised mean difference (SMD) was used to assess how similar the distribution of covariates was between remote and face-to-face consultations. A 10% difference was considered large enough to be noteworthy. For statistical disclosure reasons, summary statistics were suppressed in groups with fewer than 10 consultations.

### Statistical methods

#### Choice of estimator

We used targeted maximum likelihood estimation (TMLE) to estimate the difference in prescribing rates between patients seen remotely and those seen face-to-face. TMLE is a doubly robust method that involves modelling both the outcome and the treatment mechanism using ensemble models while still yielding valid standard errors. This is followed by a targeting step that optimizes the bias-variance trade-off for the parameter being estimated. It uses cross-validation to minimise overfitting.

#### Choice of algorithm

We included a generalized linear model (GLM), generalized additive model, random forest, xgboost, Multivariate Adaptive Regression Splines and lasso net GLM as learners for both our treatment and outcome ensemble model. Using a discrete super learner – which compares the ensemble model with the individual learners – to pick the final model was not computationally feasible so the ensemble model was used with non-negative linear least squares as the metalearner. As this has been proven to perform at least as well as any individual model asymptotically, we considered this a reasonable simplification. The analysis was carried out using R 4.0.2 using sl3 and tmle3 for the modelling.

### Sensitivity Analysis

We reran the models without the mixed consultations that were included in the face-to-face group in the main analysis.

### Role of the funding source

No external funding

## Results

There were 45,997 consultations for ARIs (34,555 unique patients), of which 61% (28,127) were remote and 39% (17,870) face-to-face. For children, 48% of consultations were remote whereas for adults 66% were remote. Antibiotics were prescribed in 48% of all consultations for adults, and in 52% of remote and 42% of face-to-face consultations. For children, 43% of all consultations led to antibiotic prescriptions with 42% in remote and 43% in face-to-face consultations (Table 1). The median age was 4 in children (0-16) and 49 for adults (16+). In children, 52% of consultations were male compared to only 38% in adults. Ethnicity was missing for a higher proportion of consultations for children than for adults. Adult consultations were evenly distributed across the IMD quintiles but there was a slight over-representation of child consultations in the most deprived IMD quintile (Table 1).

**Table 1.**
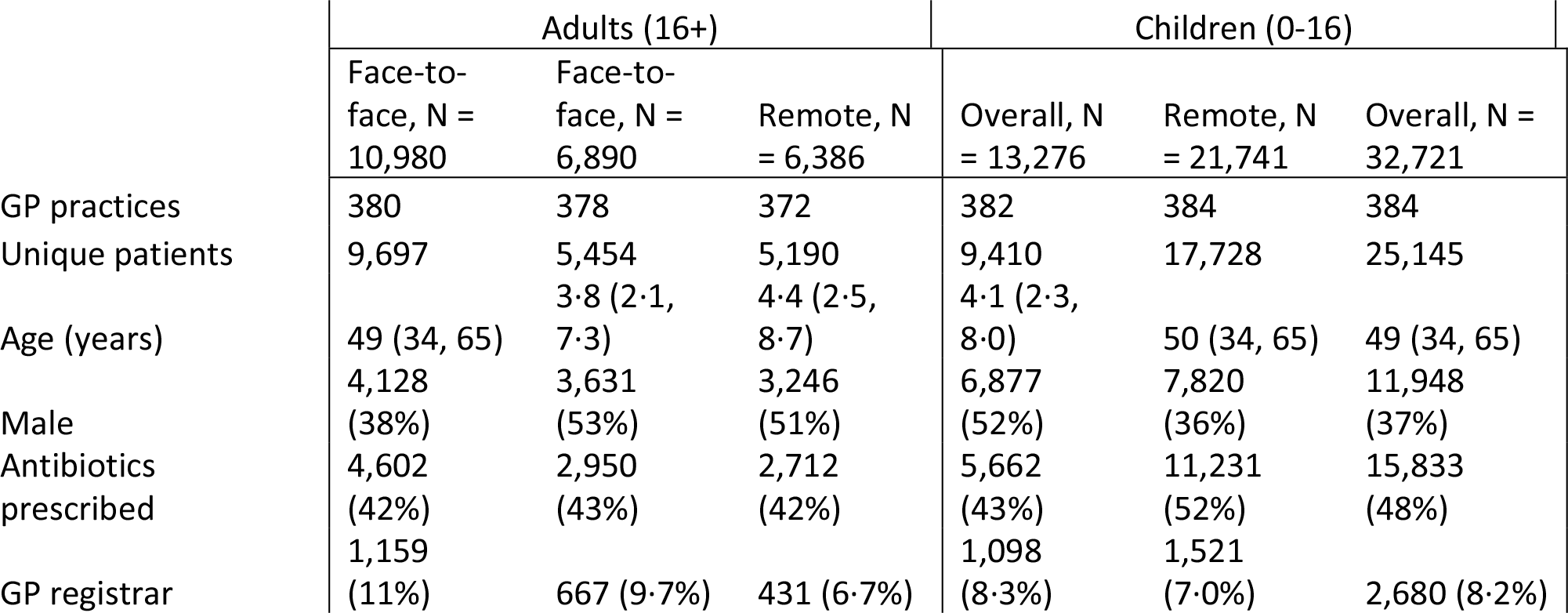
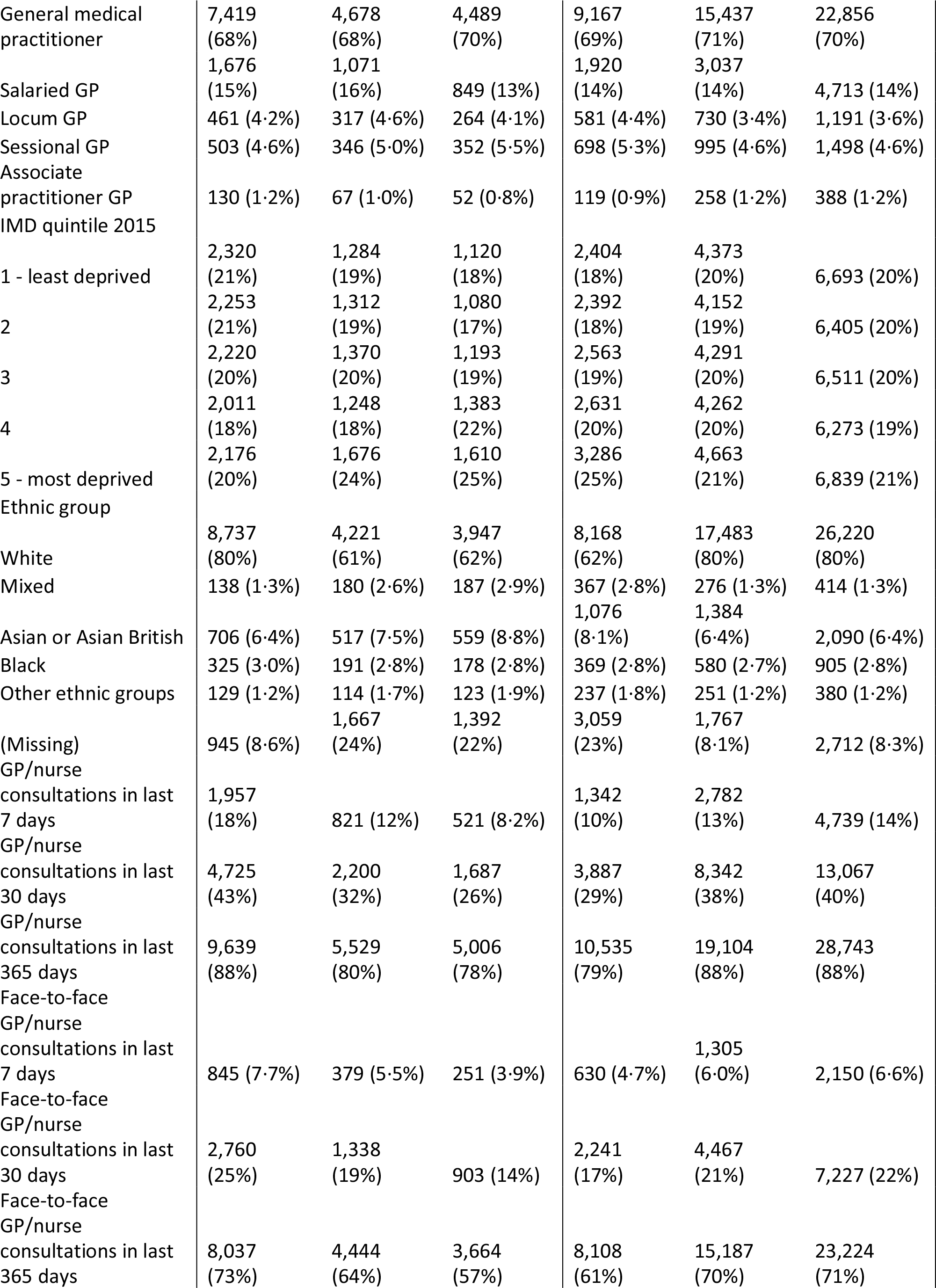
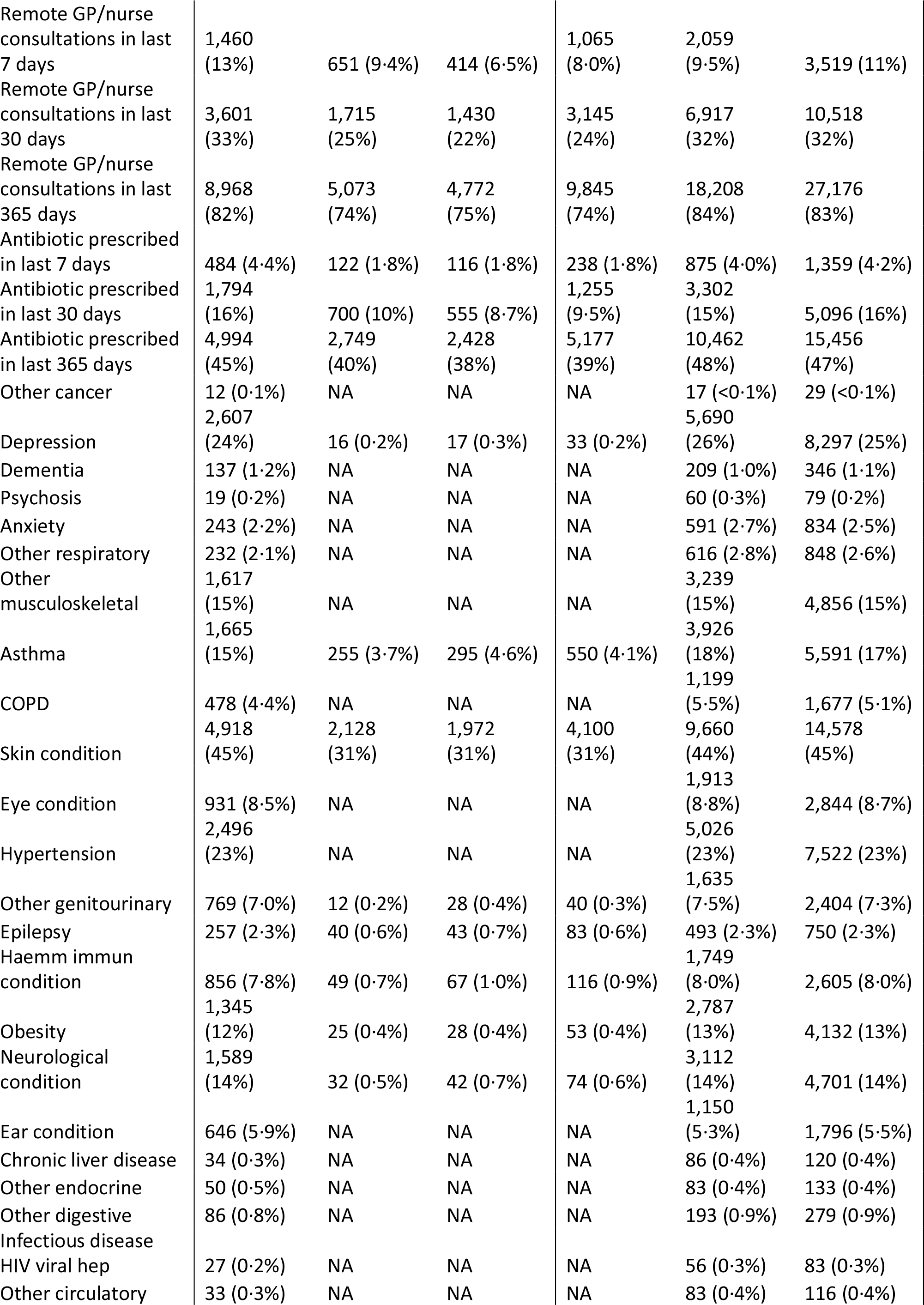
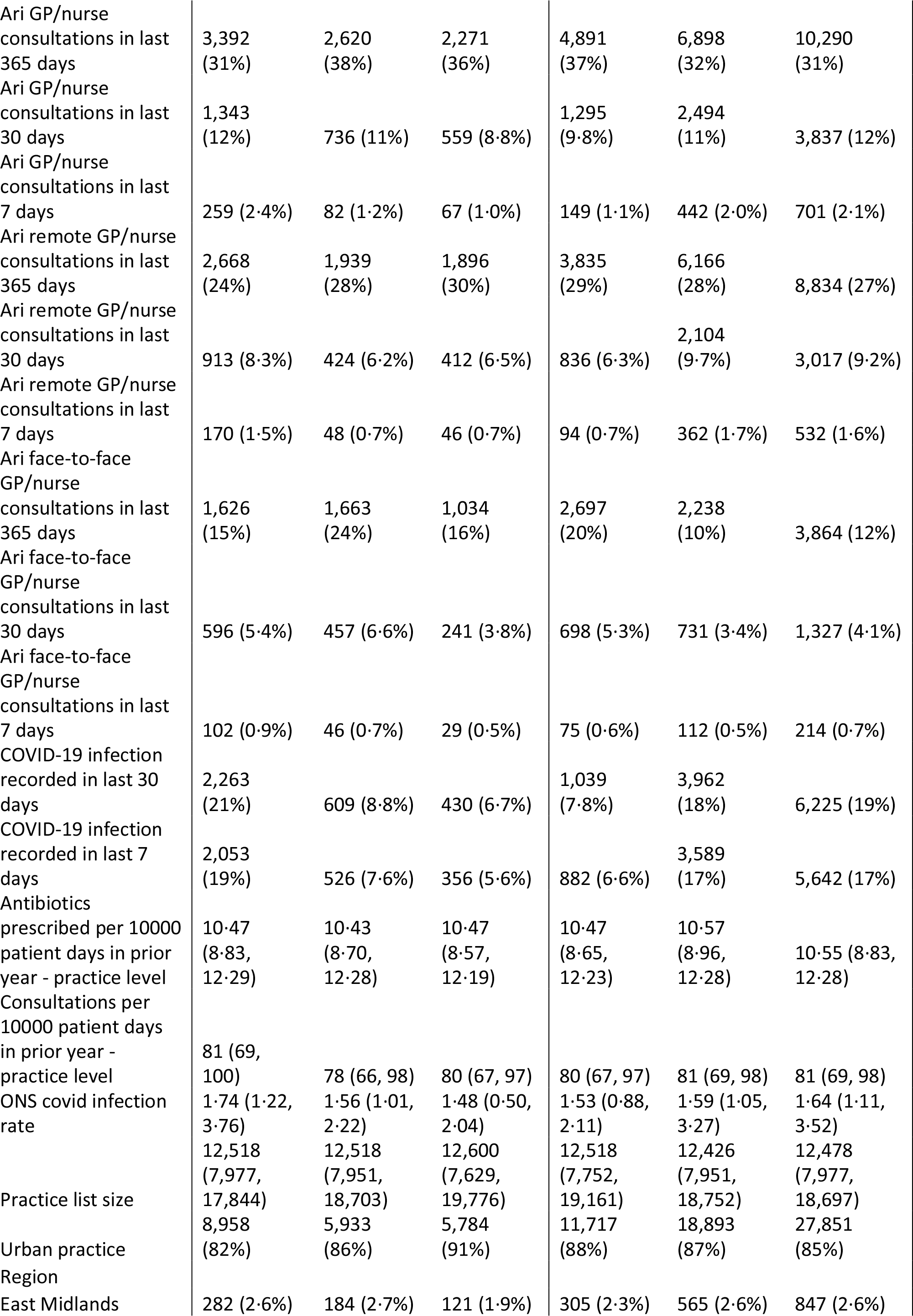
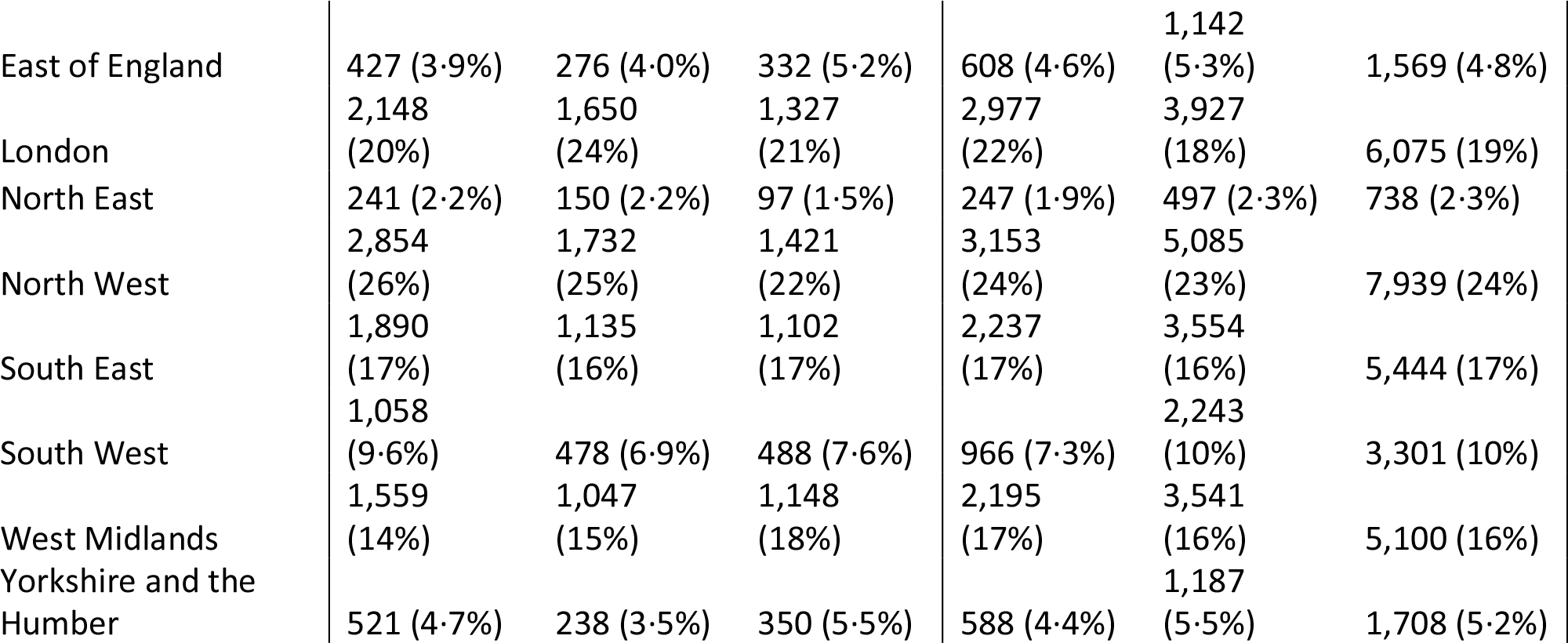
Baseline characteristics based on consultations as the unit of analysis

Regardless of the type of infection, adults had a higher proportion of remote consultations compared to children. For a URTI, 71% of adults were seen remotely compared with just 53% of children (Figure S1). The most common ARI type for both adults and children was URTI (49·8% and 70·0% of all diagnoses respectively) but the frequency of some infection types differed substantially between adults and children, notably LRTI being high in adults but low in children (14·3% and 5·8% respectively) and otitis media being the opposite (2·8% in adults and 10·1% in children) (Figure 1A and 1B). LRTI and otitis media had the highest antibiotic prescribing rates within the study (LRTI: 82·6% adults and 78·9% children; Otitis media: 73·2% adults and 84·3% children) so the prevalence of these infections will highly influence baseline prescribing rates. Antibiotics were more commonly prescribed for adults with URTIs in remote rather than face-to-face consultations (53·3% compared to 46·1%) whereas there was very little difference in prescribing rates for children (39·6% compared to 37·9%). Antibiotics were more likely to be prescribed in remote consultations for COVID infection for both adults (17·5% vs 7·8%) and children (15·0% vs 2·7%) (Figure 1C and 1D).

**Figure 1.**
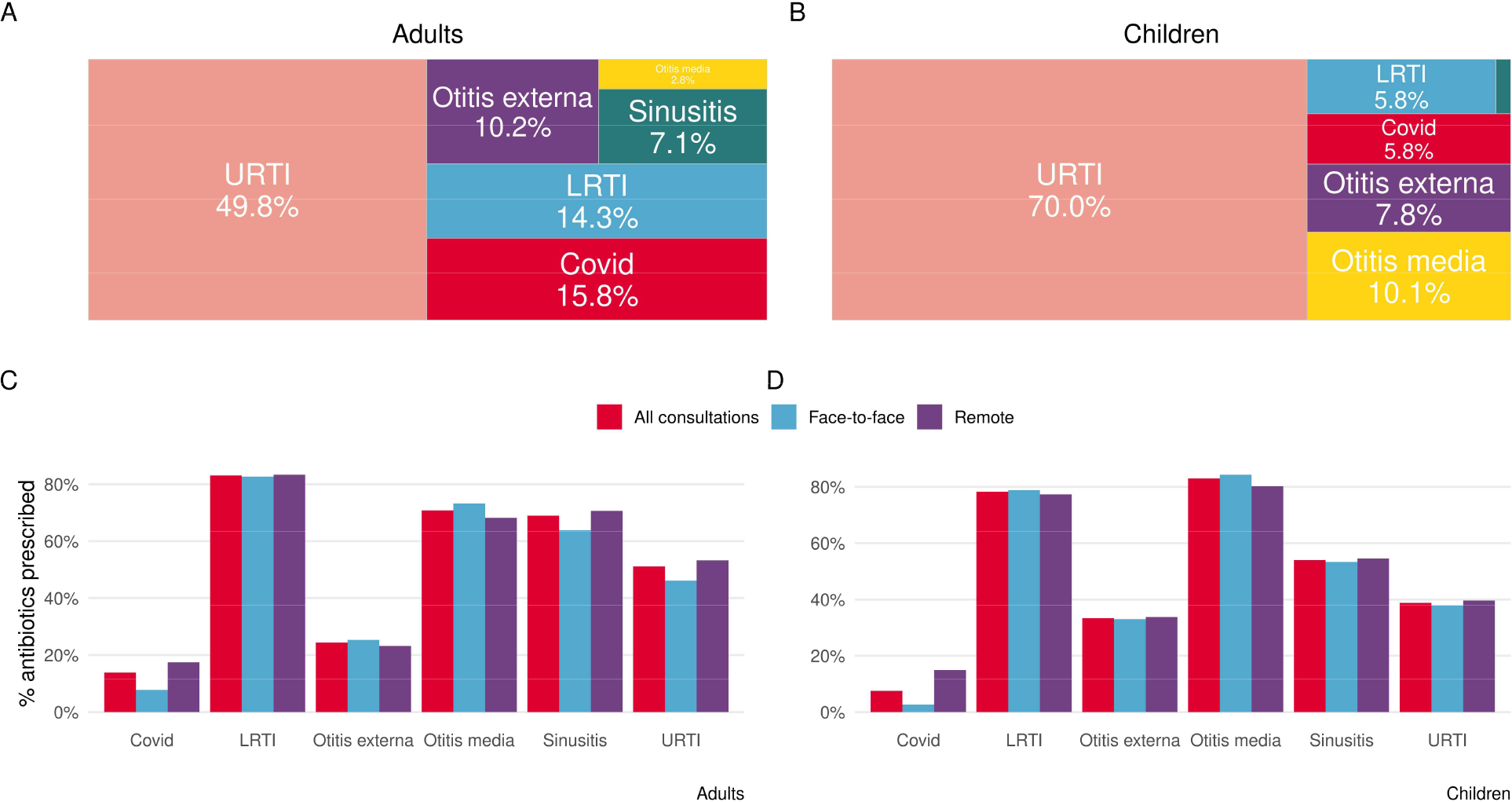
Breakdown of infection type codes (% of total) (A & B) and proportion of consultations leading to an antibiotic prescription by consultation mode and infection type (C & D)

There were some differences in the baseline characteristics of patients having remote compared with face-to-face consultations (Table 1, Figure 2). For both adults and children, consultations were more likely to be remote when related to a URTI or in an urban practice and more likely to be face-to-face when related to an otitis media diagnosis or with a GP registrar. The number of previous ARI face-to-face consultations in the last year was also associated with being seen face-to-face.

**Figure 2a.**
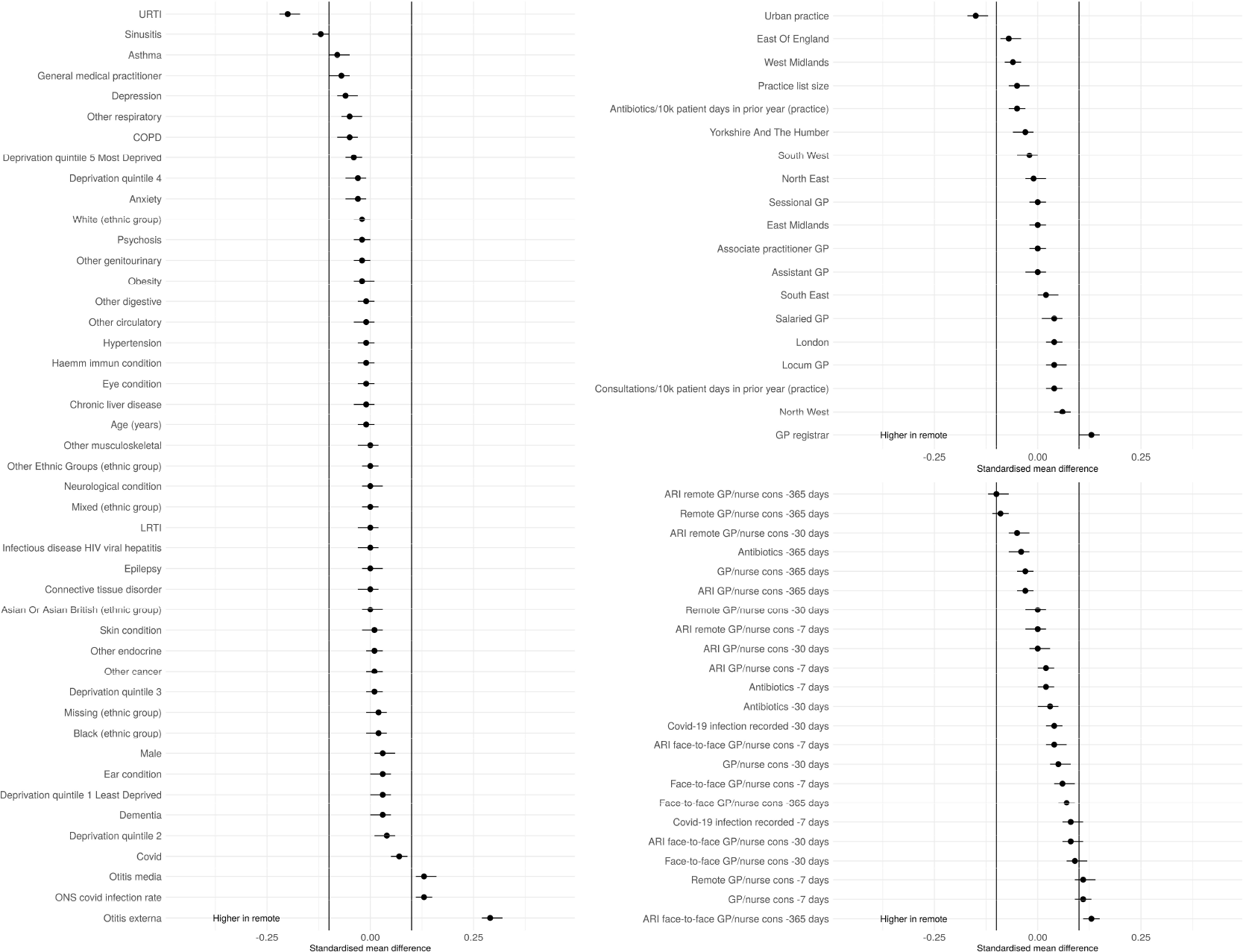
SMD adults

**Figure 2b.**
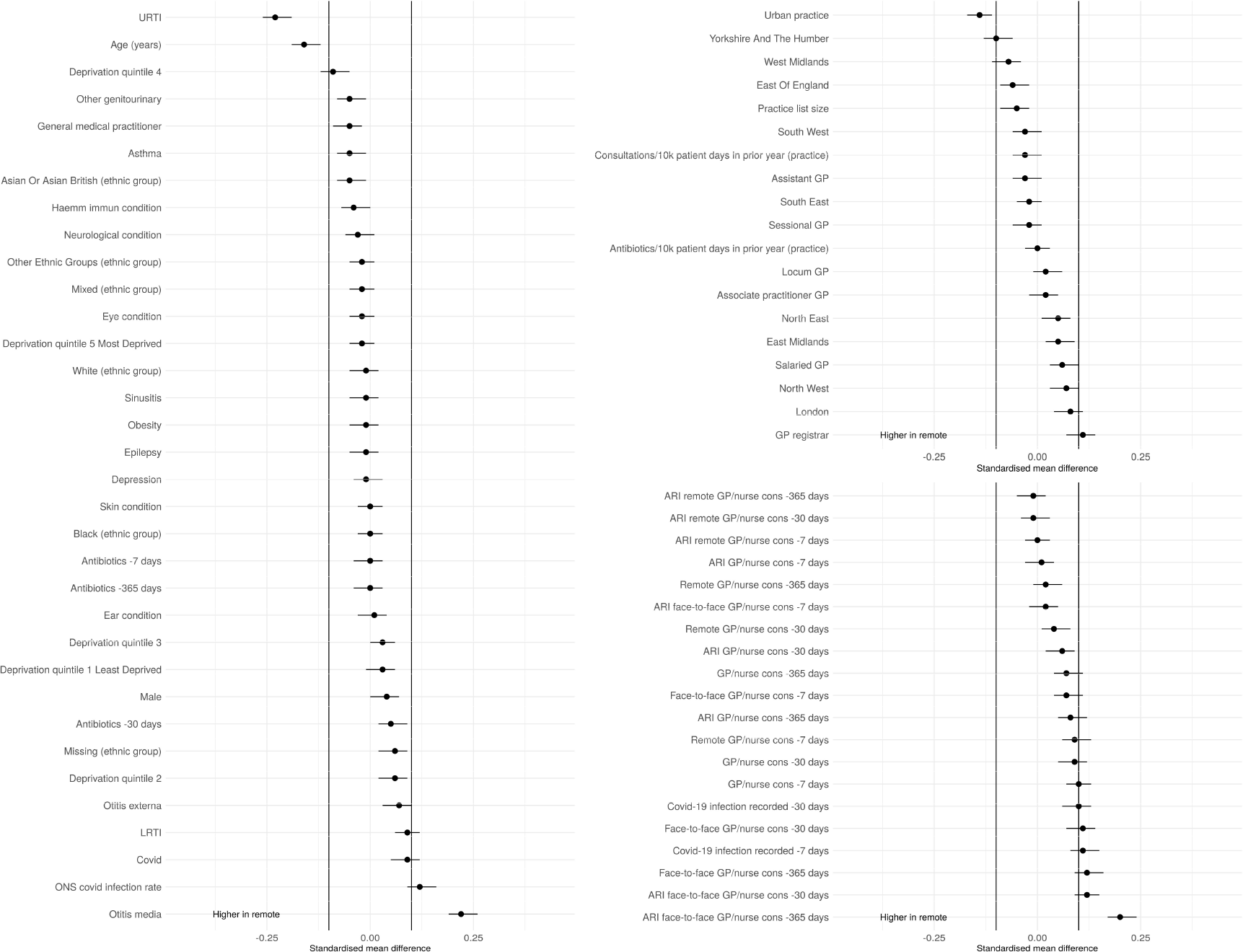
SMD children

### Treatment and outcome models

The minimum and the maximum propensity scores were 0·037 and 0·944, respectively, for children; and 0·039 and 0·962, respectively, for adults which suggests the positivity assumption is reasonable.

We estimated that 44·7% (95% CI: 43·7, 45·7) of adults would have been prescribed antibiotics if they had been seen face-to-face with 49·9% (95% CI: 49·2, 50·2) if seen remotely, which corresponds to a difference in average treatment effect of 5·1% (95% CI: 4·0, 6·2) and an odds ratio of 1·23 (95% CI: 1·18, 1·29) after adjustment using TMLE.

In children, we estimated that 41·8% (95% CI: 40·6, 43·0) would have been prescribed antibiotics if they had been seen face-to-face with 42·8% (95% CI: 41·6, 43·9) if seen remotely. This corresponds to a non-significant difference in average treatment effect of 1% (95% CI: -0·5, 2·6) and an odds ratio of 1·04 (95% CI: 0·98, 1·11)

#### Sensitivity Analysis

The results without mixed mode consultations are consistent with the main analysis (Supplementary S2).

## Discussion

Using data from almost 46,000 GP consultations for ARIs in general practice in England we estimated differences in antibiotic prescribing between remote and face-to-face ARI consultations. After adjusting for a variety of factors, we found that an adult who had a remote consultation was 23% more likely to be prescribed antibiotics compared to if they had had a face-to-face consultation. In contrast, we found no evidence of an association between consultation mode and antibiotic prescribing in children.

The overall antibiotic prescribing rates by infection type are broadly similar to previously published estimates. (25,26) However, this study adds new insight on the use of remote consultation for different infection sub-groups. Remote consultations are used for all types of ARIs, but there is variation in the proportion of remote consultations by infection type. Patients are most likely to be seen remotely for sinusitis, and least likely to be seen remotely for otitis media. As the need for antibiotics also differs by infection type, the unadjusted baseline prescribing rate for remote and face-to-face consultations are different. This shows the importance of interpreting unadjusted estimates cautiously.

As the existing evidence on remote consultations and antibiotic prescribing is mixed (9–12), our finding that antibiotics are more likely to be prescribed to adults in remote rather than face to face consultations represents an important contribution to the field. We applied TMLE to a large patient-level dataset with a wide range of variables. The rich data means that we can control for more demographic, socioeconomic and clinical variables than most studies and the machine learning methods allow for more complex non-linear relationships between variables to be accounted for. Therefore, this provides the strongest evidence yet that antibiotic prescribing is higher in remote ARI consultations compared to face-to-face for adults.

We do not observe a difference by consultation mode in antibiotic prescribing for children. This could mean that there is no difference or that we were unable to detect it. Previous studies have found a difference in children but in different settings. (15) The decision to prescribe depends on many factors including comorbidities and acuity. As children often have few comorbidities, acuity and other unobserved factors would be more important when prescribing antibiotics to children so there is more potential for unobserved confounding.

Including both adults and children in the same study provides us with a unique opportunity to compare results from the same practices during the same time period. A potential explanation for why we see a difference in adults but not in children could be that GPs are more risk averse when consulting with children and prefer to bring them in for a face-to-face consultation before prescribing. We do observe a higher proportion of both face-to-face and mixed consultations in children.

The factors affecting antibiotic prescribing for ARIs, and the interaction with consultation mode are complex and will require further research to unpick. Both patients and GPs might behave differently in a face-to-face compared with a remote consultation. Total triage (patients are remotely assessed before booking into a consultation) should ensure that patients have the right type of consultation for their concern, but this system is not used by all practices, and is not perfect, especially when there is high demand for appointments. Patients may exert particular pressure on GPs in certain types of appointments. Clinical examinations, which may help to determine the need for antibiotics – such as listening to a chest or looking in an ear – are not possible in a remote consultation, which may influence prescribing. Increased GP workload has also been associated with increased prescribing of broad-spectrum antibiotics. (27)

### Implications for clinicians and policymakers

There are implications for both antibiotic prescribing and the use of remote consultations. Increased prescribing in adults could have a substantial impact on the UK’s commitment to reduce antibiotic prescribing by 15% by 2024, given ∼70% of antibiotics prescribing happens in primary care and that ARIs are the most common condition that antibiotics are prescribed for. (1,28) Antibiotic prescribing declined from 2015-2019 but data from the pandemic is harder to interpret. (29) Both patients and health care professionals have an important role to play in ensuring sustainable use of antibiotics.

Clinical guidelines should be adapted to make sure antibiotic prescribing advice for GPs factors in remote consultations. For example, many clinical risk scores used to guide antibiotic prescribing were developed for use in face-to-face consultations, so it may be necessary for separate risk scores to be developed for use in remote consultations. It is also important to consider that remote consultations have the positive externality of not requiring unwell patients to travel to the GP surgery thereby reducing the spread of respiratory infections.

There should be continued focus on educating the public on the importance of responsible use of antibiotics. Some antimicrobial stewardship activities such as the Treat Antibiotics Responsibly, Guidance, Education and Tools initiative have been adapted to work better for both prescribers and patients in remote consultations. (1)

### Future research

The risks and benefits of remote consultations in general practice are not fully understood. Further research is required to understand differences in quality of care and outcomes between remote and face-to-face consultations across a range of clinical scenarios. This study raises a concern that antibiotic prescribing rates for adults are significantly higher in remote consultations, but we do not know whether this is clinically appropriate. More work is needed to explore the appropriateness of antibiotic prescriptions across consultation modes and in different clinical contexts. That work may in turn inform our approaches to triage, by aiding understanding of clinical indications for directing patients to one consultation type over another.

Further quantitative analysis is needed to explore whether our findings hold true for other prescribers. This is especially important given the rapid rise in numbers of nurse practitioners, pharmacists and paramedics working in general practice. Qualitative investigation of the issue, including speaking to clinicians and patients, and observing ARI consultations to explore how antibiotic prescribing plays out in practice in remote vs face-to-face environments is also needed.

### Strengths and limitations

We have used a large patient-level dataset that is representative of the English population. We controlled for patient-, clinician- and practice-level factors known to be associated with antibiotic prescribing using a doubly-robust causal machine learning method. In addition, the way we classified consultations into remote, mixed and face-to-face and then grouped mixed and face-to-face consultations together, more accurately reflects the way consultations take place in general practice in England rather than using only face-to-face consultations as the comparator.

Although we included a wide range of factors, there may still be unobserved confounding, such as urgency, acuity, or staffing levels, that could influence both consultation mode and antibiotic prescribing, but we are unable to measure them in this study. This could impact the results, but due to the large effect size it is unlikely to fully remove the relationship we observe between consultation mode and antibiotic prescribing in adults.

We are unlikely to capture all ARIs due to poor clinical coding - some studies found that only 69% of antibiotics prescribed could be linked to a specific part of the body and/or clinical condition. (30) There could be a difference in the accuracy and completeness of coding (both of symptoms and antibiotic prescribing) between remote and face-to-face consultations. This could lead to bias if poor coding was applied to a greater extent to either consultation mode. We only included ARI GP consultations, not other potential prescribers, and we did not differentiate between acute and delayed prescriptions or whether the prescriptions were guideline compliant.

## Conclusion

Our results are concerning because of the potential implications for antibiotic consumption and resistance. Further research is needed to understand the causes of increased antibiotic prescribing in remote consultations, and to determine whether the observed increase is appropriate.

On a broader level, the effect of increased remote consulting in general practice needs to be considered in the round. While the rapid introduction of remote consultations was a great success in the sense that it allowed general practice to keep functioning in the first few months of the pandemic, there may be unintended consequences that follow. Careful and thorough evaluation of interventions takes on an even greater importance when changes are instituted rapidly, as was the case with the accelerated rollout of remote consultations.

## Data Availability

We used deidentified primary care data from the Clinical Practice Research Datalink (CPRD). For more information, please visit: https://www.cprd.com/Data-access, and enquiries can be emailed to. Scientific approval for this study was given by the CPRD Independent Scientific Advisory Committee (ISAC). The study was approved by the ISAC for CPRD research (20_143). The data are provided by patients and collected by the NHS as part of their care and support. The primary care data can be requested via application to the Clinical Practice Research Datalink.

## Acknowledgments

We would like to thank Joe Standing (UCL GOS Institute of Child Health) and Adam Steventon (Our Future Health) for discussions about the design and method. We also thank Eleanor Coster and Rebecca Fisher for help with the interpretation of the findings from a GP perspective. Finally, we want to thank Arne Wolters (The Health Foundation) for his support. The IAU works in partnership with NHS England and we would like to thank Minal Bakhai and Dr Jean Ledger from NHS England for their support and advice while progressing this work.

## Footnotes

## Author contributions

EV and GC conceived the study. EV and KDC developed the study design and analytical approach, in consultation with other project team members. EC processed the data sources. EV and KDC performed the statistical analyses. EV, KDC and PC drafted the initial version of the manuscript. All authors contributed to the interpretation of the findings and reviewed and edited the manuscript for intellectual content. All authors approved the final version of the manuscript and agreed to be accountable for all aspects of the work. EV is the guarantor. The corresponding author attests that all listed authors meet authorship criteria and no others meeting criteria have been omitted.

## Funding

No external funding was received for this work.

## Ethical considerations

CPRD has ethics approval from the Health Research Authority to support research using anonymised patient data. Requests by researchers to access the data are reviewed via the CPRD Research Data Governance (RDG) and the protocol number for this study is : 21_000357. The RDG process is to ensure that the proposed research is of benefit to patients and public health. The data used in this study were obtained from practices that had consented to participate in CPRD research, and all data were handled in accordance with CPRD guidelines for data management and confidentiality.

## Conflicts of interest

No conflicts of interest.

## Notes

### Competing Interest Statement

The authors have declared no competing interest.

