## Supplementary Materials for "Antibiotic prescribing in remote versus face-to-face consultations for acute respiratory infections in English primary care: An observational study using TMLE"

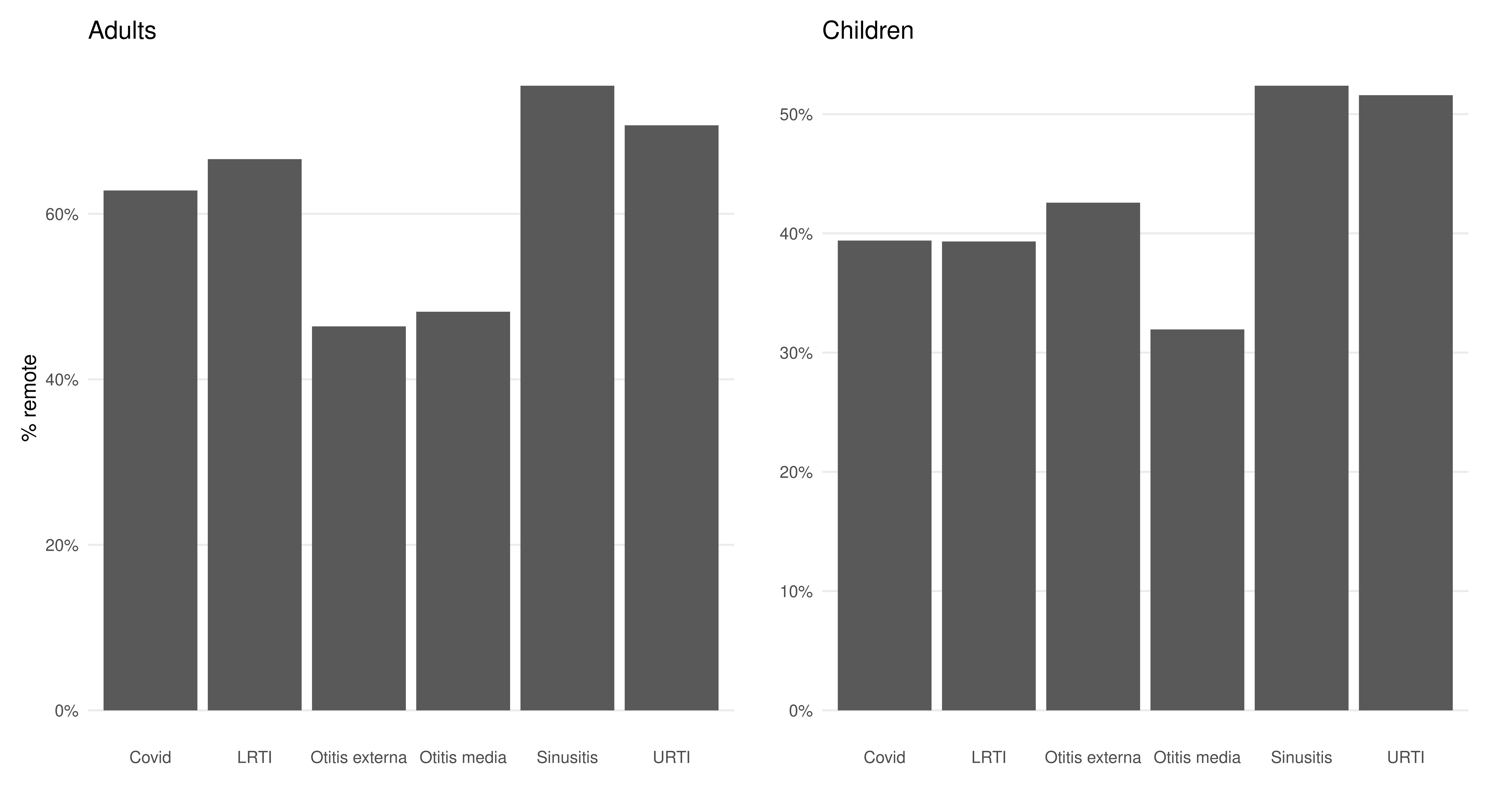

Figure S1 Percent of consultations that were remote by infection type

S1. Link to codelists to identify ARIs.

https://github.com/THF-evaluative-analytics/antibiotic-prescribing-cprd/tree/main/Codelists

S2. Outcome model excluding mixed consultations

We estimated that 44.2% (95% CI: 43.1, 45.3) of adults would have been prescribed antibiotics if they had been seen face-to-face with 50.0% (95% CI: 49.3, 50.6) if seen remotely, which corresponds to a difference in average treatment effect of 5.7% (95% CI: 4.5, 6.9) and an odds ratio of 1.26 (95% CI: 1.20, 1.32) after adjustment for a variety of demographic, clinical and socio-economic factors using TMLE.

In children, we estimated that 42.7% (95% CI: 41.2, 44.2) would have been prescribed antibiotics if they had been seen face-to-face with 43.0% (95% CI: 41.8, 44.9) if seen remotely. This corresponds to a non-significant difference in average treatment effect of 0.5% (95% CI: -1.4, 2.3) and an odds ratio of 1.01 (95% CI: 0.94, 1.09)
